# Prosthetic Visual Acuity with the PRIMA System in Patients with Atrophic Age-related Macular Degeneration at 4 years follow-up

**DOI:** 10.1101/2023.11.12.23298227

**Authors:** MMK. Muqit, Y. Le Mer, L Olmos de Koo, FG Holz, JA Sahel, D. Palanker

## Abstract

**Objective:** To assess the efficacy and safety of the PRIMA subretinal neurostimulation system 48-months post-implantation for improving visual acuity (VA) in patients with geographic atrophy (GA) due to age-related macular degeneration (AMD) at 48-months post-implantation.

**Design:** First-in-human clinical trial of the PRIMA subretinal prosthesis in patients with atrophic AMD, measuring best-corrected ETDRS VA (Clinicaltrials.gov NCT03333954).

**Subjects:** Five patients with GA, no foveal light perception and VA of logMAR 1.3 to 1.7 in their worse-seeing “study” eye.

**Methods:** In patients implanted with a subretinal photovoltaic neurostimulation array containing 378 pixels of 100 μm in size, the VA was measured with and without the PRIMA system using ETDRS charts at 1 meter. The system’s external components: augmented reality glasses and pocket computer, provide image processing capabilities, including zoom.

**Main Outcome Measures:** VA using ETDRS charts with and without the system. Light sensitivity in the central visual field, as measured by Octopus perimetry. Anatomical outcomes demonstrated by fundus photography and optical coherence tomography up to 48-months post-implantation.

**Results:** All five subjects met the primary endpoint of light perception elicited by the implant in the scotoma area. In one patient the implant was incorrectly inserted into the choroid. One subject died 18-months post-implantation due to study-unrelated reason. ETDRS VA results for the remaining three subjects are reported herein. Without zoom, VA closely matched the pixel size of the implant: 1.17 ± 0.13 pixels, corresponding to mean logMAR 1.39, or Snellen 20/500, ranging from 20/438 to 20/565. Using zoom at 48 months, subjects improved their VA by 32 ETDRS letters versus baseline (SE 5.1) 95% CI[13.4,49.9], p<0.0001. Natural peripheral visual function in the treated eye did not decline after surgery compared to the fellow eye (p=0.08) during the 48 months follow-up period.

**Conclusions:** Subretinal implantation of PRIMA in subjects with GA suffering from profound vision loss due to AMD is feasible and well tolerated, with no reduction of natural peripheral vision up to 48-months. Using prosthetic central vision through photovoltaic neurostimulation, patients reliably recognized letters and sequences of letters,and with zoom it provided a clinically meaningful improvement in VA of up to eight ETDRS lines.

## Introduction

Age-related macular degeneration (AMD) is a leading cause of irreversible vision loss associated with increasing age^1^. It was projected to affect 196 million people by 2020^2^. The late and advanced forms of AMD, macular neovascularization (MNV) and geographic atrophy (GA), are associated with severe visual impairment^1–3^ and affect 1.49% of the US population above 40 years of age, corresponding to a prevalence rate of approximately 0.94%^3^.

Geographic atrophy, which affects approximately 8 million people worldwide^4^, is associated with a gradual loss of photoreceptors in the macula, encompassing the center, which is responsible for high-resolution vision. This can severely impair visual functions, such as reading and face recognition. Low-resolution peripheral vision is retained in this condition, necessitating the use of eccentric fixation. Therefore, any treatment strategy to provide functional central vision should not jeopardize the surrounding healthy retina.

Although photoreceptors are lost within geographic atrophy, the inner retinal neurons largely survive^5^. PRIMA is a wireless prosthesis in which photovoltaic pixels directly convert projected light into patterns of electric current^6,7^ to reintroduce visual information into the degenerate retina by electrical stimulation of second-order neurons—the bipolar cells.

Preclinical studies in rodents have demonstrated that such stimulation results in a network-mediated retinal response, which preserves many features of normal vision: flicker fusion at high frequencies (>20 Hz)^8,9^ with adaptation to static images^10^, “on” and “off” responses to increments and decrements in light with antagonistic center-surround^10^, and non-linear summation of the inputs from bipolar cells into ganglion cells’ receptive fields (called sub- units)^11^, essential for high spatial resolution.

The first clinical version of this implant (PRIMA, Pixium Vision, Paris, France) is 2 mm wide (corresponding to about 7° of the visual angle in a human eye) and 30 μm thick, containing 378 pixels of 100 µm in width (**Figure 1**). Images captured by the camera mounted on agmented-reality glasses are processed and projected onto the retina and implant using near-infrared (880 nm) light to avoid photophobic and phototoxic effects of bright illumination^24^ (**Figures 2**). Current flowing through the retina between the active and return electrodes in front of the pixels stimulates the nearby inner retinal neurons^12^, which then pass the responses to ganglion cells, thereby harnessing residual retinal signal processing^9,13,14^. To avoid irreversible electrochemical reactions at the electrode–electrolyte interface, stimulation is pulsed and charge-balanced^15^. For a steady perception under pulsed illumination, sufficiently high frequencies (30 Hz) are applied to enable flicker fusion.

**Figure 1:**
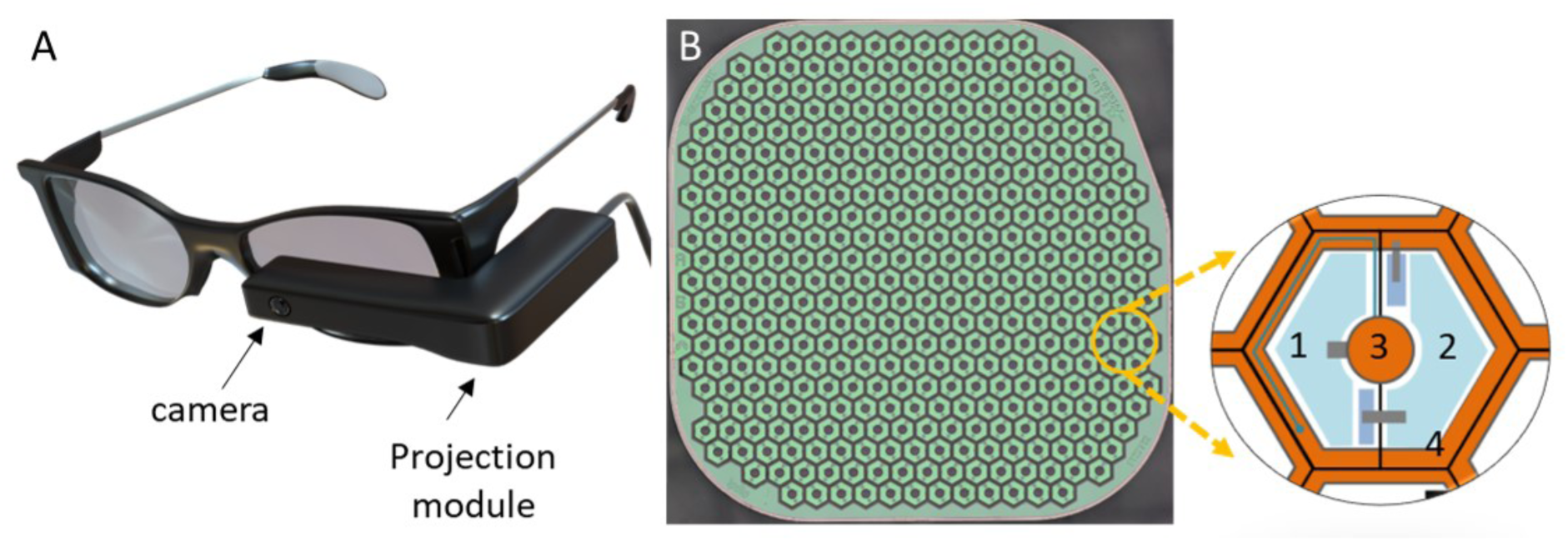
PRIMA system and the implant. A) Transparent augmented reality glasses PRIMA-2. B) Implant of 2x2mm in width and 30µm in thickness is composed of 100µm wide hexagonal pixels, which include 2 photodiodes (#1,2) connected between the central active electrode (#3) and a circumferential return electrode (#4).

**Figure 2:**
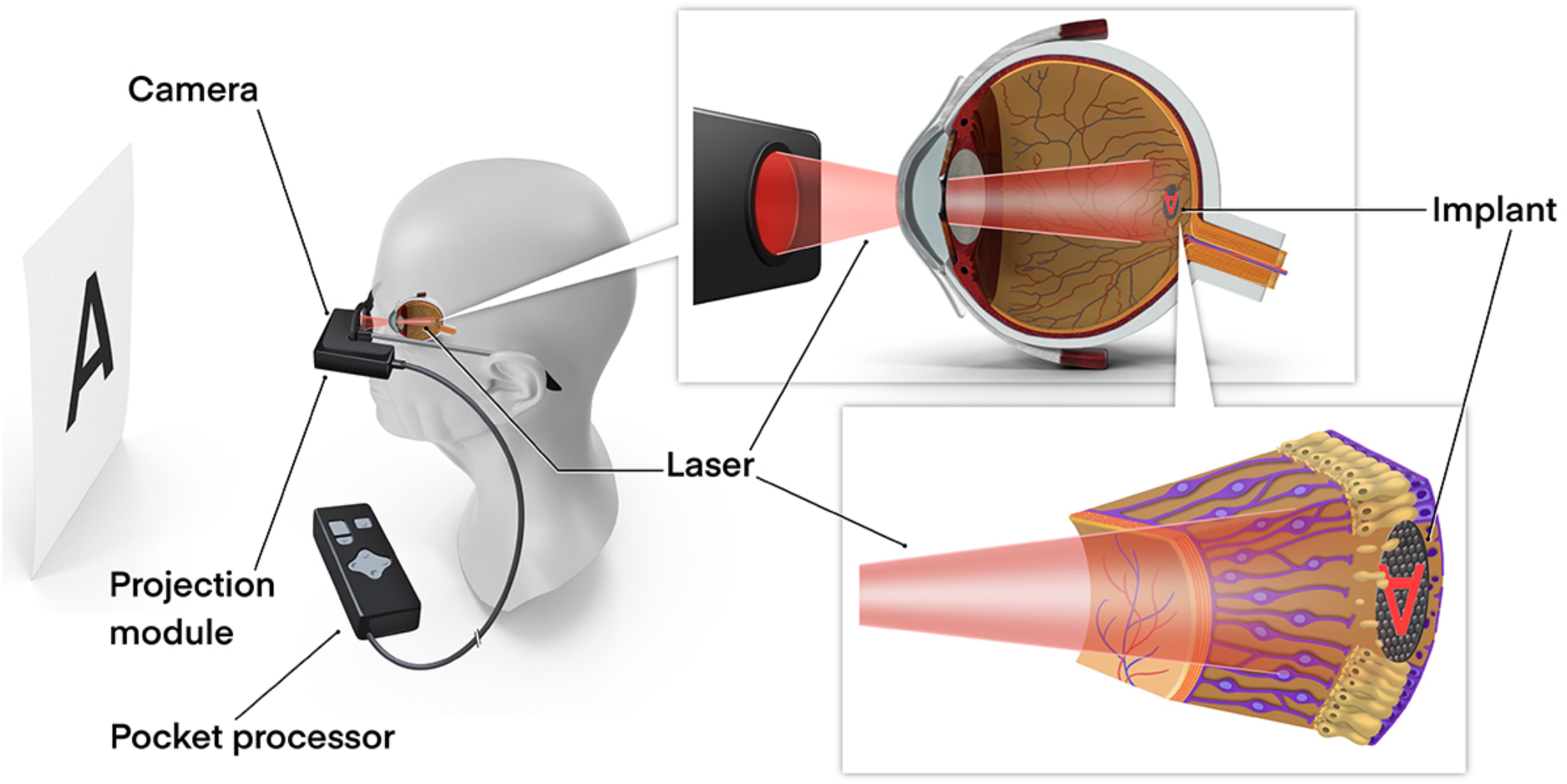
Simplified diagram of the PRIMA-2 system. Photovoltaic implant in subretinal space in one eye. Images captured by a camera are projected into the eye from augmented reality glasses connected to a pocket image processor.

The electric field is highly localized because of the presence of the active and return electrodes in each pixel^9^: preclinical testing in rodents with 75 µm and 55 µm pixels demonstrated grating acuity matching the pixel pitch^9,16^. Perceptual tests of the PRIMA implants with 100 μm pixels in non-human primates demonstrated similar stimulation thresholds to those observed in rodents and responses (saccadic movement) down to a single pixel activation^17^.

To assess prosthetic vision independently of the remaining natural vision in the first phase of the study, opaque video glasses with a digital mirror display were used. The initial primary efficacy endpoint was light perception at the implant location, assessed by an Octopus 900 perimeter (Haag-Streit, Switzerland). The 12-month results were reported in 2020, demonstrating that sub-macular implantation of the PRIMA array in patients with geographic atrophy is feasible, with no decrease in eccentric natural acuity, eliciting visual percepts in the former scotoma through neurostimulation with the PRIMA array^18^.

In October 2019, the PRIMA glasses were modified from an opaque (virtual reality) to a transparent, augmented reality (AR) design (Figure 1A). We demonstrated that patients could use their natural peripheral vision in combination with central prosthetic vision ^19^. A secondary endpoint was added to the study: measurement of visual acuity (VA) between 18 and 24 months using Landolt C optotypes. VA was measured in the implanted eye with and without PRIMA- 2 glasses, with and without zoom, allowing a magnification of up to ×8. To distinguish prosthetic vision from the residual natural one, contrast in prosthetic vision was inverted, so that subjects typically perceive white letters on a black background. In 2022, we reported the 24-month follow-up results, demonstrating implant stability and that Landolt C acuity without zoom closely matched the 100 μm pixel pitch: 1.17 ± 0.13 pixels, corresponding to the average of logMAR 1.39, or 20/500 on a Snellen scale, ranging from 20/438 to 20/565.^19^. We also reported that thickness of the retinal layers remained stable during the follow-up of 36 months, with no adverse structural abnormality across the implant^20^.

In 2022 an ETDRS test of VA was added to the protocol as a secondary endpoint. Here, we report the safety profile and prosthetic vision achieved under these settings at the 4-year follow-up.

## Methods

### Patients

This feasibility study of the PRIMA implant (NCT03333954) aimed to test safety and functionality in five patients with atrophic AMD. The study adhered to the Declaration of Helsinki and received ethical approval from the Comité de Protection des Personnes Ile de France II and the Agence Nationale de Sécurité du Médicament et des Produits de Santé. Study participants were above 60 years of age and had advanced dry AMD with an atrophic zone of at least three optic disc diameters and best-corrected VA of ≤20/400 in the worse-seeing study eye; no foveal light perception (absolute scotoma) but visual perception in the periphery, with preferred retinal locus determined by microperimetry; absence of photoreceptors and presence of the inner retina in the atrophic area, as confirmed by optical coherence tomography (OCT); absence of macular neovascularization (MNV) verified by retinal angiography. All other ocular and general pathologies that could contribute to low VA were excluded. Patients provided written informed consent to participate in the study. VA in the non-implanted fellow eye was measured throughout the study as a control. Surgical procedures were performed in the Fondation Ophtalmologique A. de Rothschild (Paris, France), with the first subject implanted in December 2017 and the fifth in June 2018^20^. The patients’ rehabilitation and visual function assessment were carried out at the Clinical Investigation Center of the Quinze-Vingts National Eye Hospital (Paris, France).

### PRIMA implantation

For the implantation surgery, the surgeon (YLM) examined the fundus image and OCT scans in a pre-planning phase. Based on the preferred retinal locus (PRL) positions and atrophy size, the PRIMA implantation site was verified. Retinotomy location was selected based on pre- operative fundus image to reduce the risk of jeopardizing the useful residual vision, including the PRL. A complete vitrectomy was completed, with removal of the posterior hyaloid membrane. The retina was detached between the macula and the planned position of the retinotomy by injecting balanced salt solution to form a bleb. The detached retina was marked with diathermy, and then incised on a length of 3mm at 4 to 5 mm distance from the fovea. A subretinal spatula was used to detach the macula, with care taken to avoid button-holing any thinned areas of the macula, and avoiding formation of a full-thickness macular hole. During surgery, the PRIMA implant was delivered through the retinotomy and into the subretinal space at the macula using silicone-tipped forceps. A bubble of perfluorocarbon liquid was injected over the macula to flatten the fluid bleb and stabilise the PRIMA implant at the submacular docking site. After the PRIMA implant was docked in the subretinal space, a pick was used to move the chip into the optimal location.

### Assessment of natural visual acuity

The best-corrected VA of each patient without the PRIMA glasses was measured to assess any potential VA loss after implantation. ETDRS letter charts were used at 4 meters; if the patient could not correctly identify at least 20 letters at 4 meters, the test was continued at 1 meter. The smallest font size which the patient could read at least four letters in one line was recorded.

### Assessment of prosthetic vision

A camera on the AR PRIMA-2 AR glasses captured a visual field of 50° × 40°, with a central third of the image being projected onto the retina to match the display’s 17° field of view without zoom. The projected images covered a field of 5.1 mm (17°) on the retina, with a resolution of 10.5 µm. Maximum peak retinal irradiance was 3.5 mW/mm^2^, with maximum pulse duration of 10ms and frame rate of 30 Hz, well within the thermal safety limits for chronic use of near-infrared light^21^. Computer-generated images could also be projected on the ocular display directly, independently of the camera. The angular magnification of the system without zoom was 1:1. In the measurements of the letter recognition, patients were also allowed to use the electronic zoom at their preferred level of magnification (×1, ×2, ×4 or ×8), and this setting was recorded.

VA measurements at 48 months using ETDRS charts were performed up to three times on different days near the 48-month follow-up. Results are reported as the median values of these three measurements. The ETDRS chart was placed 1 meter from the subject and the subject was asked to read as many letters as possible from top to bottom. The baseline VA test without PRIMA glasses was also performed at 1 meter, but only once because at the time it had not been defined as an efficacy endpoint.

### Assessment of safety

Investigators reported all the adverse events. These were categorized as serious or non-serious, and whether they were related to the procedure or a device. During follow-up, investigator assessments were reviewed by a clinical event committee.

### Data synthesis and analysis

To estimate the mean visual acuity within study settings, a mixed regression model with random effect for subjects was used.^22^ This corresponds to the regression model with logMAR and ETDRS VA as output variable for natural and prosthetic vision respectively, study setting as input variable, and random effects for subjects. The estimates are reported as mean standard error (SE) for the mean, 95% Confidence Intervals (CI) for the mean, and as mean differences SE for the mean difference, and 95% CI for the mean difference. The estimated means were than compared between study settings using post hoc Tukey method. The adjusted p value less than 0.05 was considered statistically significant. The analysis was performed using R version 4.1.2 (R Core Team; Vienna, Austria 2021).^23^

To test changes within study and non-study eyes as well to compare the possible differences between them, mixed regression model was developed with VA as output variable; time, eye status (study eye, non-study eye) and interaction of time with eye status as input variables; and finally with subject as random effect.

## Results

The first patient was enrolled at the Rothschild Foundation and Quinze-Vingts National Eye Hospital in November 2017 and the last in May 2018. Details of the PRIMA implantation surgery have been published earlier^18,20,24^. Analysis of VA at 4 years was possible for three of five patients. All the implants remained stable and functional in the subretinal space (**Figure 3**). Two patients were excluded (**Figure 4**): one died from cancer, unrelated to the study, with no data available after the 12-month visit, and the other received an intrachoroidal implantation in error.

**Figure 3:**
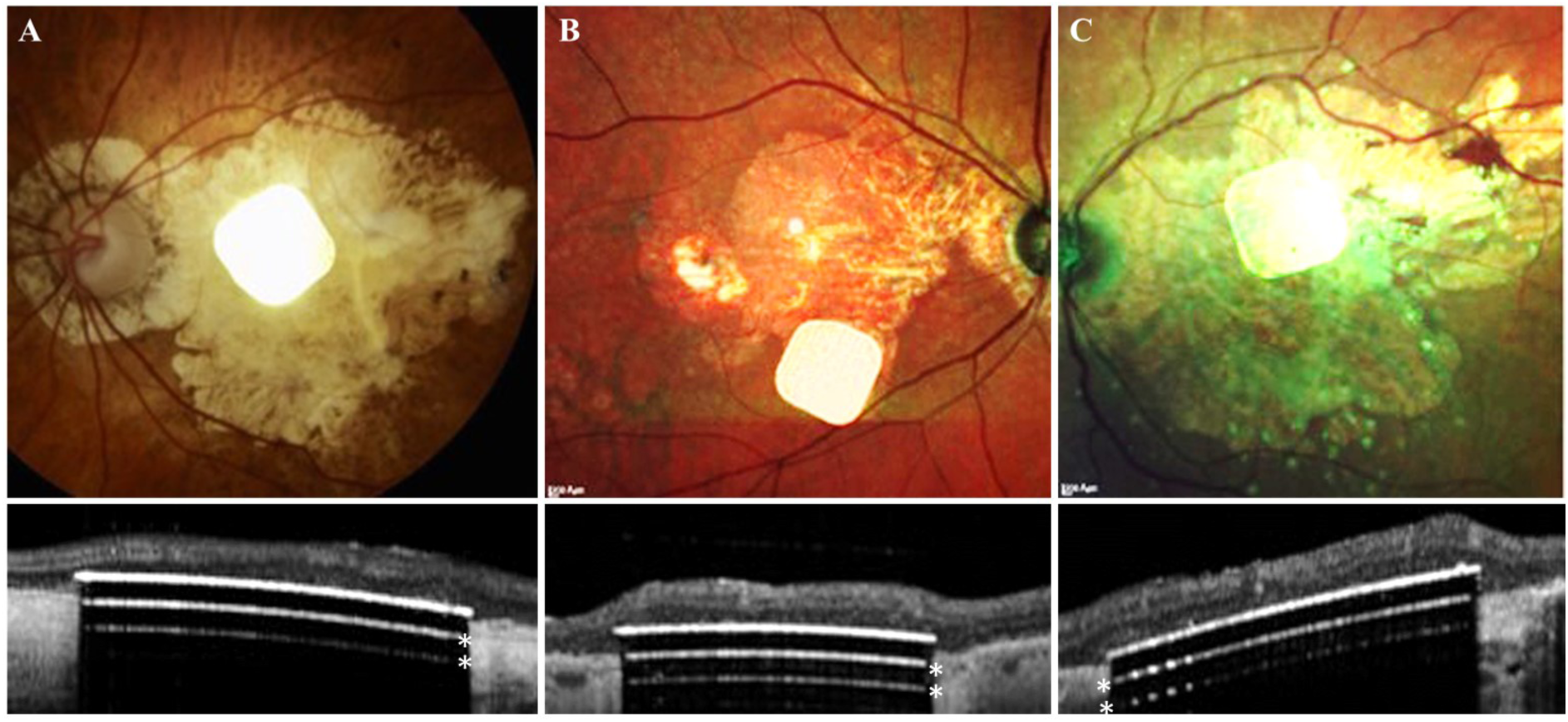
Fundus photographs and OCT of the subretinal implants at 48-month in three patients (A, B, and C). Two white lines below the implant surface (*) are the OCT artifacts due to strong light reflection from the implant surface.

**Figure 4:**
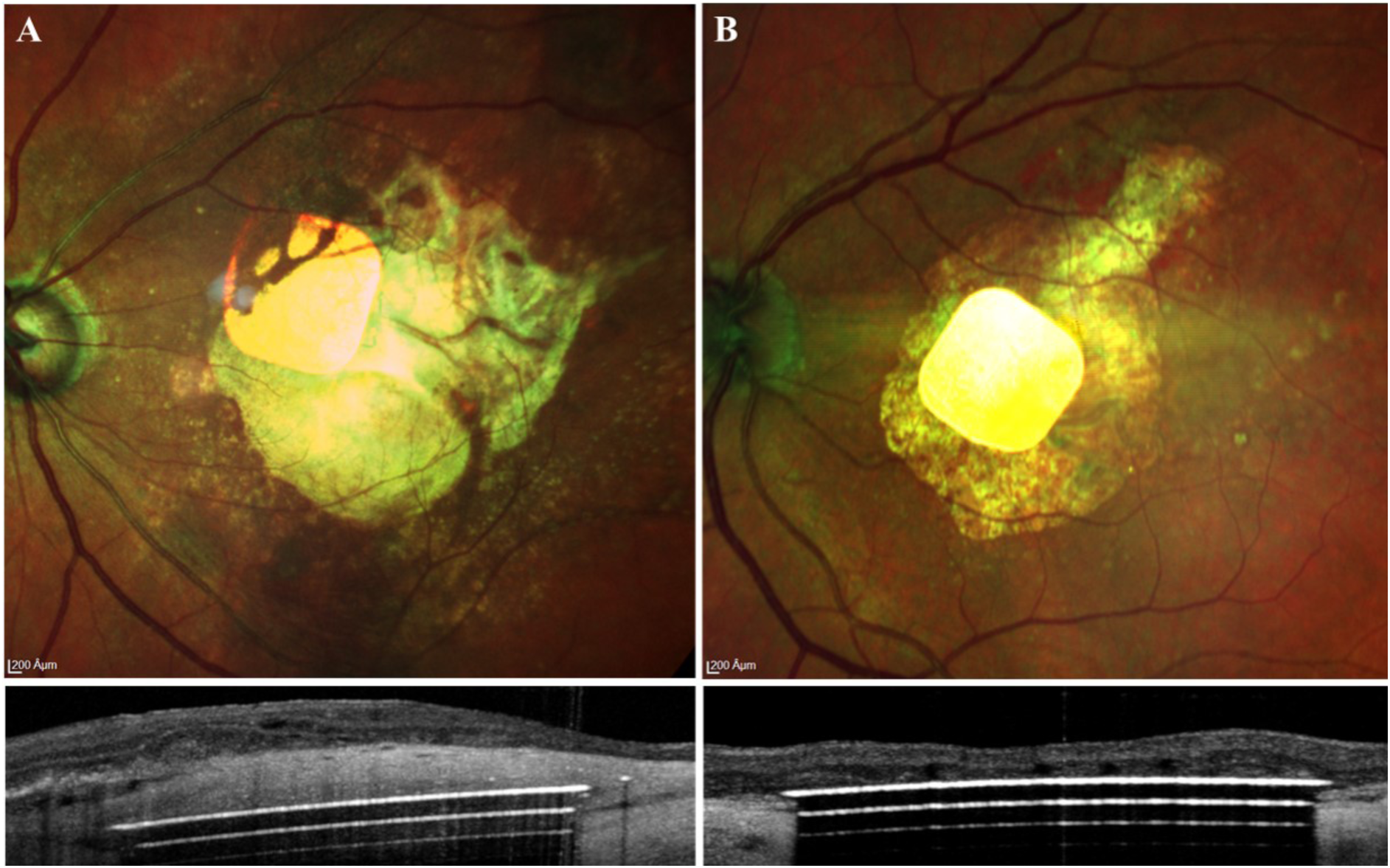
Fundus photographs and OCT of the excluded patients. 48-month fundus image (first row), and 48-month OCT scan (second row). (A) Implant in the unintended location (inside choroid). (B) Implant at 12-months in a deceased patient.

### Natural Vision

During the 48-months follow-up, all subjects experienced slight reduction of VA in the non-study eye, albeit not significant. For all 5 patients, the mean baseline VA in the non-study eyes was logMAR 0.78 (SE 0.13) 95%CI [0.41,1.15], and in the study eyes, it was logMAR 1.48 (SE 0.13) 95%CI [1.11, 1.85], p<0.001. At the 48-month visit, in the non-study eyes, the estimated mean VA was logMAR 1.03 (SE 0.14) 95% CI [0.63,1.42]; in the study eyes, the mean VA was logMAR 1.33 (SE 0.14) 95% CI [0.93, 1.72], p=0.058. Residual natural VA in the treated eye of any patient did not decrease during the 48-month follow-up period. It even exhibited a temporary improvement, albeit not significant: p=0.801 (**Figure 5**). By 48 months, the mean VA in the study eyes improved by 0.1 logMAR mainly due to one subject who improved by up to logMAR 0.4, while in other subjects it improved by no more than logMAR 0.1.

**Figure 5:**
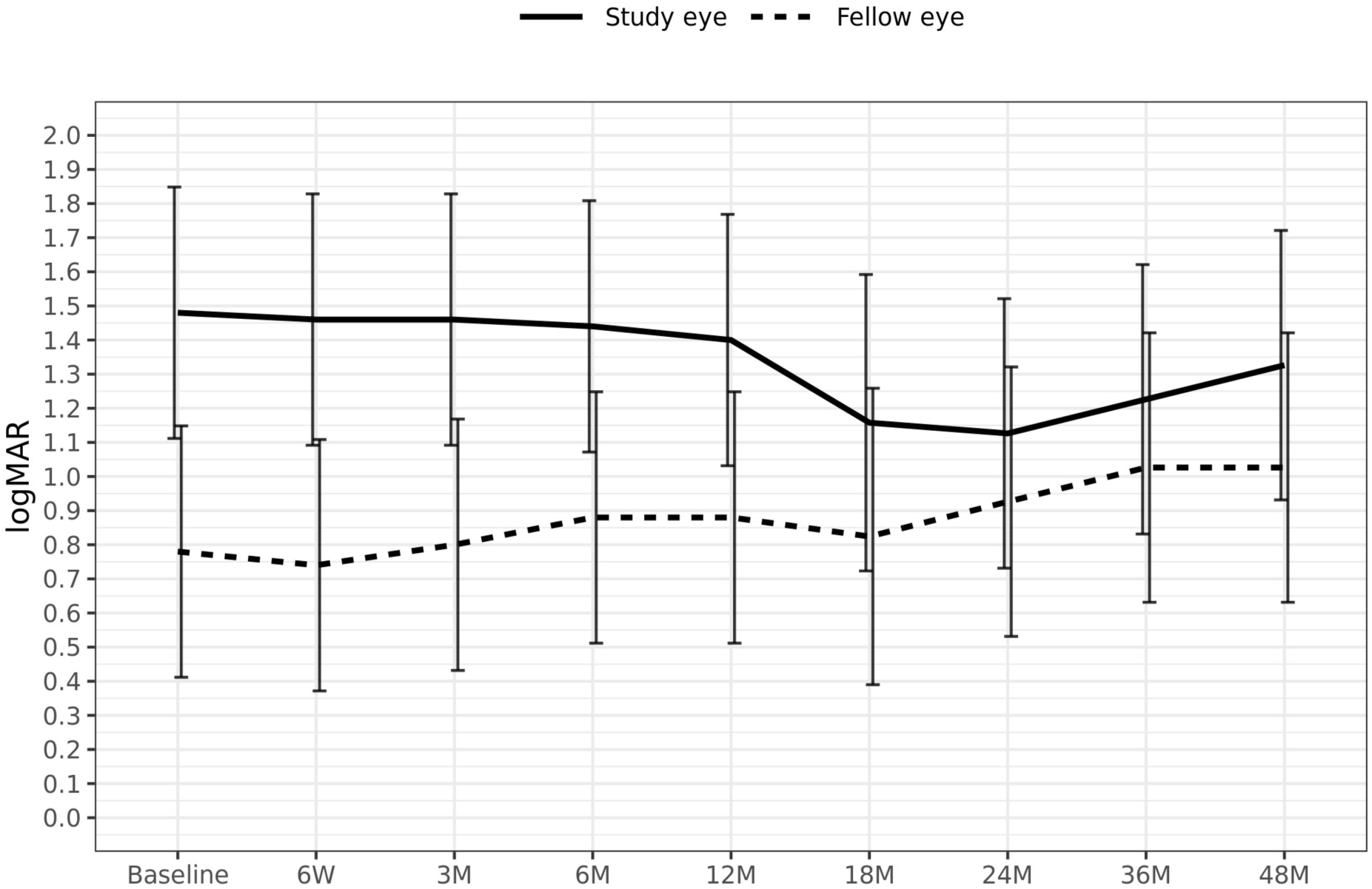
Mean natural visual acuity over time for all patients. Error bars represent confidence intervals

### Prosthetic vision

The PRIMA system inverted the contrast of the ETDRS chart, so that subjects perceive the black letters on a white board as bright stimulation patterns on a black background. As shown in **Figure 6**, VA improved in all three subjects compared to baseline, and their prosthetic vision (with the PRIMA glasses) was better than natural vision (without PRIMA glasses). The means for each settings are: Baseline: 11.7 ETDRS letters (SE 4.8) 95% CI [0, 32.1]; without PRIMA glasses at 48 months: 18.2 ETDRS letters (SE 4.8) 95% CI[0,38.6]; and with PRIMA glasses (prosthetic vision with preferred magnification) at 48 months: 43.3 ETDRS letters (SE 4.8) 95% CI [22.9,63.8].

**Figure 6:**
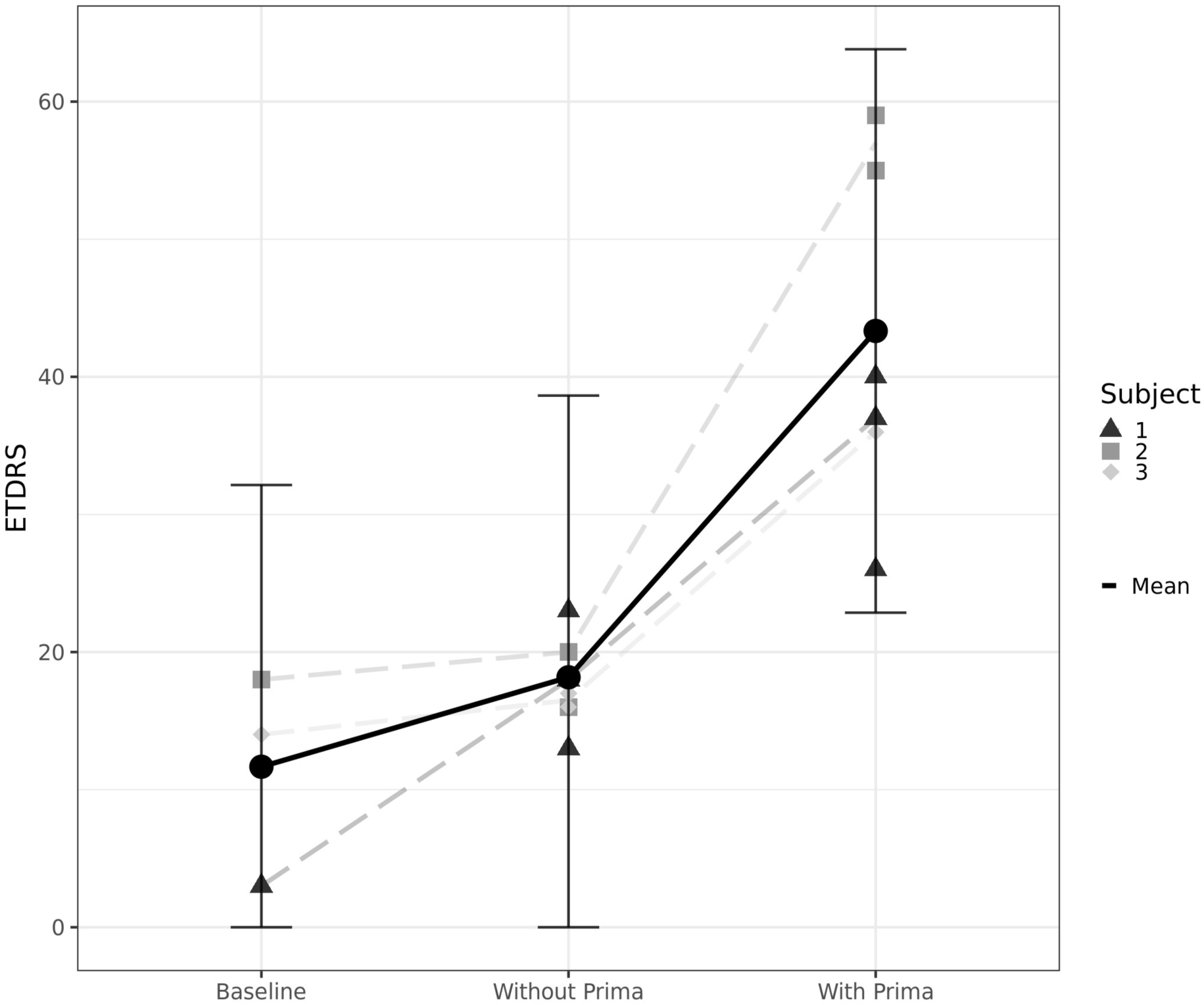
Median best-corrected prosthetic VA with the ETDRS chart at 1 meter distance.

The differences between the settings were: natural vision at 48 months (without PRIMA glasses) compared to baseline: +6.5 ETDRS letters (SE 5.1) 95% CI [-11.7,24.7], p=0.480; prosthetic vision (with PRIMA glasses) compared to baseline: +31.7 ETDRS letters (SE 5.1) 95% CI [13.4,49.9], p=0.008; and prosthetic vision compared to natural vision at 48 months (with-PRIMA versus without-PRIMA glasses): +25.2 ETDRS letters (SE 5.1) 95% CI [6.9,43.4], p=0.017. Both differences, between prosthetic vision and baseline, as well as between prosthetic vision and natural vision at 48 months, are significant.

In addition to objective VA measurements, subjects were observed during training sessions. Given their ability to recognize letters, they were able to read sequences of letters and words. This was also observed during exercises, like reading product names on food packages, reading panels outdoor and indoor, or reading a train timetable (supplementary video).

### Safety

In total, four study related serious adverse events (SAEs) were reported in the follow-up of all five patients, but none was related to the device (**Table 1**). Per investigator assessment, one event was definitely procedure related, two were probably procedure related and one was possibly procedure related, as described below. The subject’s death due to cancer was neither procedure nor device related.

**Table 1:** Procedure-related serious adverse events in all five patients.

| Adverse Event Term | Number of Patients |
| --- | --- |
| Ocular hypertension | 2 |
| Macular neovascularization | 1 |
| Retinal detachment | 1 |

The MNV event, that was deemed to be procedure-related, was observed in an area where Bruch’s membrane got damaged during implantation surgery under local anesthesia. An unexpected head movement of the subject at the time of retinal implant delivery under the macula led to damage of the Bruch’s membrane and choroidal hemorrhage. After resorption of blood, fibrotic tissue was observed around the implant. This patient had visual perception of stimulation, but no spatial resolution using the implant, and hence this patient was excluded from the acuity measurements (Figure 3A). 31 months after implantation, an asymptomatic MNV was discovered in the same eye of this subject, within an area that was not touched during surgery. Therefore, it was judged as unlikely to be procedure- or device-related. The MNV was treated with a course of intravitreal ranibizumab injections and resolved after a year without sequelae. At 42 months’ post-implantation, the same subject developed a second MNV in a different area of the posterior pole near the retinotomy site. This second MNV event was deemed as procedure-related.

12 months post-op, a retinal detachment developed in the control eye of another patient. This was successfully treated with vitrectomy and gas injection, and the retina was re-attached. This subject was also known to have underlying glaucoma at baseline, and developed uncontrolled ocular hypertension in the implanted eye 2 years post-op, which was unresponsive to medical treatment. The patient underwent successful trabeculectomy with eye pressure control. Since vitrectomy risks exacerbation of the pre-existing glaucoma, this adverse event was deemed procedure-related, even though the patient already had glaucoma.

All the study-related non-serious adverse events are listed in **Table 2**. Microcysts were observed in three subjects, which resolved without sequelae. The microcysts did not create any symptoms and seemed not to affect perception of the stimulation by the implant. Two patients developed significant ocular hypertension that required ongoing medical treatment.

**Table 2:** Study related non-serious adverse events and the number of affected patients.

| Adverse Event name | Patients [n] |
| --- | --- |
| Macular microcyst | 3 |
| Ocular hypertension | 2 |
| Submacular perfluorocarbon liquid | 1 |
| Choroidal hemorrhage | 1 |
| Localized subretinal bleeding | 1 |
| Corneal epithelial defect | 1 |
| Retained silicone oil droplet | 1 |
| Punctuate keratitis | 1 |
| Headache | 1 |
| Myodesopsia | 1 |
| Ocular discomfort | 1 |
| Peripheral retinal tear | 1 |
| Choroidal detachment in periphery | 1 |
| Vitreous hemorrhage | 1 |
| Subretinal chip displacement | 1 |
| Choroidal neovascularization | 1 |

Implant stability in the subretinal space has been previously reported.^20^ The authors have also published data that showed that in the 3 patients reported in this study, the PRIMA implants did not cause any obvious structural changes in the vicinity of the device, nor in the outer retinal layers over a 3-year period.^20^

## Discussion

Up to 4 years after implantation, the subretinal photovoltaic implant activated by PRIMA-2 glasses enabled three subjects to read at least four additional lines on the vision chart compared with their natural vision. There were some adverse events (serious or not), which were managed efficiently. This did not bring into question the safety and biocompatibility of the device or the patients’ ability to tolerate the PRIMA implant.

A key feature of the PRIMA subretinal photovoltaic system is the ability of implanted subjects to consistently recognize letters and to follow a sequence of letters. In this study, all subjects read more letters with PRIMA-2 glasses than without. The difference between prosthetic vision with zoom versus baseline was 32 letters (range of 22 letters to 39 letters). At the 4-year time- point, the mean gain was 25 letters, which corresponds to logMAR 0.5 (five lines). Even without zoom, PRIMA system compares favorably with the previous epiretinal and subretinal prosthetics^25–29^. With zoom, it demonstrated much higher letter acuity, albeit on the account of the correspondingly reduced visual field. Such visual gains are clinically meaningful for patients with foveal geographic atrophy secondary to AMD.

The function of zoom for advanced AMD patients is offered by two other technologies: the implanted telescope, such as IMT (VisionCare Ophthalmic Technologies, Saratoga, USA)^30^ and Virtual Reality glasses, such as eSight (eSight Eyewear, Toronto, Canada)^31^. These two devices utilize PRL and peripheral retina and therefore preclude normal use of peripheral vision. So far, neither of these two approaches has gained much popularity. PRIMA, on the other hand, enables normal use of residual peripheral vision, while providing central prosthetic vision with optional zoom.

One of the major visual disabilities for patients with advanced AMD and, in particular, with geographic atrophy is the gradual decline and then a permanent loss of reading ability within the central field.^32–34^ Unlike the current and potential pharmacological treatments for geographic atrophy, which aim to slow down the growth of atrophic lesions without any functional improvement in VA^35^, our results demonstrate restoration of central vision in the former scotoma.

According to the literature, the expected vision loss due to progression of GA is up to logMAR 1.5 in 5 years.^36,37^ Surprisingly, not only the natural VA in the implanted eye did not decrease, but it temporarily improved in all four subjects. The reason for this is unknown. It could be due to a neurotrophic benefit of the subretinal surgery or of the electrical stimulation^38^, or it could be a result of the vision rehabilitation training, which may enhance the gaze control and thus improve the eccentric fixation. However, over time this effect seems to be compromised by the natural degradation of patients’ vision.

The vast majority of the device- and procedure-related non-serious adverse events occurred within the first 6 months post-implantation, confirming the absence of longer-term safety issues. None of the non-serious adverse events, either device- or procedure-related or not, reasonably bring into question the overall safety of the system. These include the microcysts discovered in three subjects (two occurred 9 months post-implantation) and the asymptomatic choroidal neovascular membrane in one subject (which occurred 31 months after implantation).

In summary, the wireless photovoltaic array provided form vision with letter acuity closely matching the 100 μm pixel size in patients with geographic atrophy, with no decline in eccentric natural acuity over a 4-year period. The transparent (AR) glasses allow simultaneous use of the prosthetic central and natural peripheral vision. Using electronic zoom, patients demonstrated clinically meaningful and statistically significant improvements in vision. These results were accompanied by an acceptable safety profile. Reduced pixel size in the future implants^39,40^ may further improve the prosthetic VA to even higher levels.

## Data Availability

All data produced in the present work are contained in the manuscript

## Acknowledgements

We would like to thank the patients who participated in the study and the Pixium Vision team who designed, fabricated, and tested the PRIMA system. We are also grateful to the scientific and medical advisory board of Pixium Vision for their guidance on the clinical trial design. We would also like to acknowledge the scientific, R&D, medical, and clinical research staff who continue the patients’ care, rehabilitation, and evaluation. Finally, we acknowledge the support within the clinical event committee from Professor Andrea Cusumano and Professor Jan van Meurs.

**Supplementary Video:** Patient using PRIMA-2 system to read railway timetable information. Computer screen showing the patient view using the spectacle’s camera video projection to scan the letters using the PRIMA-2

## Notes

Financial support: This study was funded by Pixium Vision and in part by the Sight Again project under the Structural R&D Projects for Competitiveness (PSPC) and Investment for the Future (PIA) funding, managed by Bpifrance. DP was funded in part by the National Institutes of Health (grant R01-EY027786). The Biomedical Research Centre at Moorfields Eye Hospital, supported in part by the National Institute for Health and Care Research, UK, is also acknowledged. The Clinical Investigation Center at the Quinze-Vingts National Hospital is supported in part by the Inserm-DHOS, France.

Conflict of Interest: MMKM, YLM, LODK, FH, DP are consultants to Pixium Vision. JAS discloses founding shares and past consulting for Pixium Vision. DP’s patents are licensed by Stanford University to Pixium Vision.

### Competing Interest Statement

MMKM, YLM, LODK, FH, DP are consultants to Pixium Vision. 
JAS discloses founding shares and past consulting for Pixium Vision. DP patents are licensed by Stanford University to Pixium Vision.

### Clinical Trial

NCT03333954

### Funding Statement

This study was funded by Pixium Vision and in part by the Sight Again project under the Structural R&D Projects for Competitiveness (PSPC) and Investment for the Future (PIA) funding, managed by Bpifrance. DP was funded in part by the National Institutes of Health (grant R01-EY027786). The Biomedical Research Centre at Moorfields Eye Hospital, supported in part by the National Institute for Health and Care Research, UK, is also acknowledged. The Clinical Investigation Center at the Quinze-Vingts National Hospital is supported in part by the Inserm-DHOS, France.

### Author Declarations

The study received ethical approval from the Comite de Protection des Personnes Ile de France II and the Agence Nationale de Securite du Medicament et des Produits de Santa.

